# Deep learning-based identification of necrosis and microvascular proliferation in adult diffuse gliomas from whole-slide images

**DOI:** 10.1101/2025.08.14.25333656

**Authors:** Yumeng Guo, Hanli Huang, Xing Liu, Wanjing Zou, Fufang Qiu, Yunqing Liu, Ruichao Chai, Tao Jiang, Jiguang Wang

## Abstract

For adult diffuse gliomas (ADGs), most grading can be achieved through molecular subtyping, retaining only two key histopathological features for high-grade glioma (HGG): necrosis (NEC) and microvascular proliferation (MVP). We developed a deep learning (DL) framework to automatically identify and characterize these features. We trained patch-level models to detect and quantify NEC and MVP using a dataset that employed active learning, incorporating patches from 621 whole-slide images (WSIs) from the Chinese Glioma Genome Atlas (CGGA). Utilizing trained patch-level models, we effectively integrated the predicted outcomes and positions of individual patches within WSIs from The Cancer Genome Atlas (TCGA) cohort to form datasets. Subsequently, we introduced a patient-level model, named PLNet (Probability Localization Network), which was trained on these datasets to facilitate patient diagnosis. We also explored the subtypes of NEC and MVP based on the features extracted from patch-level models with clustering process applied on all positive patches. The patient-level models demonstrated exceptional performance, achieving an AUC of 0.9968, 0.9995 and AUPRC of 0.9788, 0.9860 for NEC and MVP, respectively. Compared to pathological reports, our patient-level models achieved the accuracy of 88.05% for NEC and 90.20% for MVP, along with a sensitivity of 73.68% and 77%. When sensitivity was set at 80%, the accuracy for NEC reached 79.28% and for MVP reached 77.55%. DL models enabled more efficient and accurate histopathological image analysis which will aid traditional glioma diagnosis. Clustering-based analyses utilizing features extracted from patch-level models could further investigate the subtypes of NEC and MVP.

## Introduction

Glioma is the most common histological type of primary Central Nervous System (CNS) cancer, with an annual incidence of 5-6 cases per 100,000 individuals, and adult diffuse gliomas account for approximately 70% of all glioma cases^1^. According to the 2021 World Health Organization (WHO) classification of CNS tumors^2^, genetic mutations and DNA methylation have been recommended as key molecular parameters to be considered for guiding clinical diagnosis, tumor grading, and therapeutic interventions. Meanwhile, histological features are also critically important during the diagnostic process. Especially the presence of necrosis (NEC) and/or microvascular proliferation (MVP) is considered as hallmarks of HGG in adult diffuse gliomas, which is more aggressive and indicates a poor prognosis^3^.

Histopathology analyses are routine assessments conducted by pathologists to microscopically examine a biopsy or surgical specimen that is processed and fixed onto glass slides^4^. Histopathology studies provide a gold standard for diagnosis, disease treatment, and prognostic evaluation. However, it has been demonstrated that for many diagnostic tasks, human assessments are highly subjective, and the reproducibility among different pathologists is less than optimal^5, 6^. For example, in the diagnosis of specific types of breast cancer, the pathological consistency can be as low as 48%^7^, a scenario also observed in prostate cancer^8^. Additionally, the examination of histopathological sections is a highly complicated and time-consuming task. Typically, it demands highly experienced doctors to meticulously examine gigapixel-sized whole-slide images (WSIs). This field necessitates more reproducible, precise, and efficient diagnostics for histopathological analysis.

The current application of deep learning in pathology analysis primarily focuses on automatic diagnosis tasks, such as tumor detection, classification and grading, image segmentation, etc. In recent years, numerous exciting breakthroughs have been achieved in this field. Earlier in 2016, the International Symposium on Biomedical Imaging (ISBI) organized a grand challenge to evaluate computational systems for the automatic detection of metastatic breast cancer in WSI of sentinel lymph node biopsies. During this challenge, a patch-level classification deep learning model achieved comparable performance to that of pathologists, showcasing the potential of deep learning to significantly enhance the accuracy of pathological diagnoses^9^. In 2019, a study implemented U-Net models (a classical CNN algorithm) for the automated multiclass segmentation of PAS-stained nephrectomy samples and transplant biopsies^10^. Previous studies have indicated that deep learning algorithms can also be used to learn the histological features from WSIs and forecast the prognosis for glioma. Mehmet and Daniel proposed a deep learning-based classification pipeline for grading glioma patients on the TCGA cohort, achieving 96% accuracy for GBM versus LGG classification and 71% accuracy for differentiating LGG WHO grade II and grade III^11^. Recently, several interesting studies have advanced the field by demonstrating that deep learning algorithms can combine histological images with genetic information to predict the clinical outcome of diseases, such as overall survival time and time to progression^12, 13, 14,15^. For example, Cooper et al. reported a computational approach that integrated histology images and genomic biomarkers information into a single unified network to predict the survival of glioma patients^16^. In 2023, Hollon et al. developed DeepGlioma, which is trained on a multimodal dataset comprising stimulated Raman histology (SRH) and genomic data, to streamline the molecular diagnosis of diffuse gliomas^17^. Wang et al. presented a CNN-based integrated diagnosis model that was capable of automatically classifying adult-type diffuse gliomas according to the 2021 WHO standard from annotated-free WSIs^18^. In 2024, Hoang et al. developed Deep lEarning from histoPathoLOgy and methYlation (DEPLOY), a deep learning model that classifies CNS tumors to ten major categories from histopathology^19^. Mahootiha et al. developed a multimodal deep learning framework using preoperative MRI and several EHR-derived clinical variables to improve postoperative recurrence prediction and risk-stratification for pediatric low-grade gliomas^20^. Jin et al. employed deep learning technologies to classify 5 major types of histopathological glioma images^21^, which greatly inspired us. They achieved 86.5% patch-level accuracy and 87.5% patient-level accuracy on the independent testing dataset. While the results were promising, they only focused on the histopathological glioma classification not the grading part which is more important for diagnosis and treatment. Our research significantly improved the automation of grading system for glioma and achieved better results at both patch and patient levels.

In this study, we developed a framework based on deep learning for the quantitatively detecting necrosis and microvascular proliferation from WSIs in glioma patients, aiming to provide objective high accuracy prediction to aid diagnosis in clinical practice (**Fig. 1, 2a, 4c**). We utilized a dataset from Beijing Tiantan Hospital to train the patch-level models namely NEC detector and MVP detector. This dataset encompassed over 600 newly collected WSIs from gliomas ranging from WHO grade 2 to grade 4 to ensure a comprehensive representation of glioma tumor grades. Initially, we trained deep learning models for necrosis and microvascular proliferation patch detection and quantification. Subsequently, we analyzed 1,699 WSIs from an independent cohort to train a patient-level model, named Probability Localization Network (PLNet). This model can be used for computer-aided patient diagnosis by integrating the patch-level predictors and spatial information features. Performance of this model was then evaluated using patient label that extracted from TCGA pathology reports. Finally, all patch-level and patient-level features were extracted from the models for further association analysis with clinical and genomic data (**Fig. 1**), highlighting the clinical significance for diagnosis and treatment.

**Figure 1.**
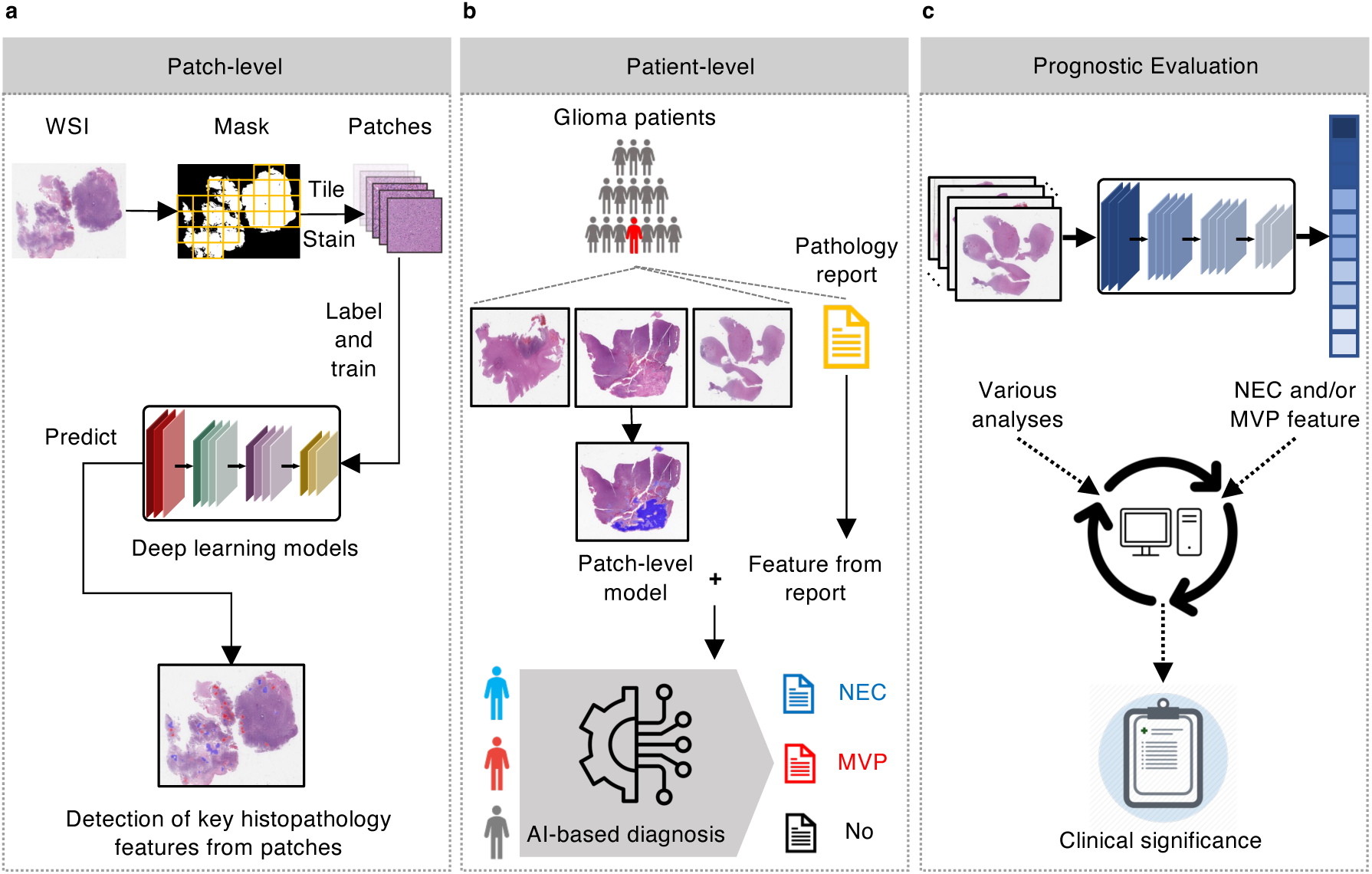
Computational and analytical pipeline of the study. (**a**) We initially applied WSI preprocessing to obtain tiled and stained patches and then developed deep learning models to detect and quantify histopathological features, including necrosis (NEC) and microvascular proliferation (MVP), at the patch level with labeled data. (**b**) We then integrated the patch-level results to aid patient-level diagnosis and grading based on an AI-based aggregation model. (**c**) Finally, we conducted several analyses, including survival analysis, differential expression analysis, and gene set enrichment analysis, to investigate the clinical implications of our computational pathology models.

**Figure 2.**
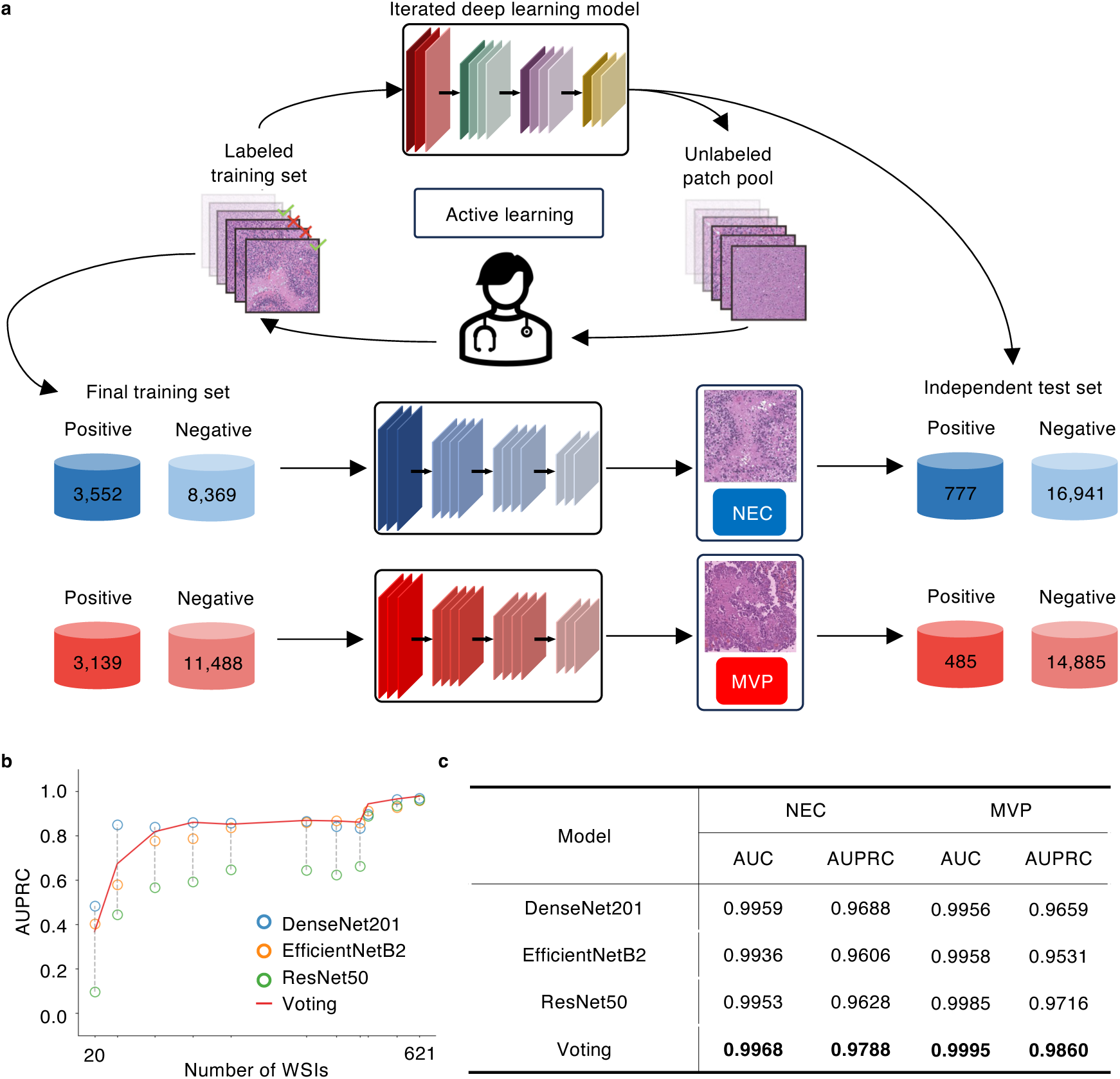
Patch level classification of necrosis (NEC) and microvascular proliferation (MVP). (**a**) Diagram summarizing the model training pipeline. An active learning strategy was applied to label patches based on whether NECs and/or MVPs were recognized in a particular patch. After active learning, the final training set included 3,552 NEC-positive, 8,369 NEC-negative patches, 3,139 MVP-positive, and 11,488 MVP-negative patches. These patches were extracted from 621 glioma WSIs. Our deep learning models were trained in the above training sets and then tested in an independent dataset, which was extracted from another 14 WSIs (777 NEC-positive, 16,941 NEC-negative, 485 MVP-positive, and 14,885 MVP-negative patches) and manually labelled by three pathologists. (**b**) Using the training set, we first trained three deep learning models using the widely used network frameworks, *i.e.*, DenseNet, EfficientNet and ResNet, and then concatenated results from these three models to generate the fourth results based on voting. Performance of the four results on test set were compared. Using necrosis-recognition model as an example, we demonstrated that the performance of patch-level models improved as the size of training dataset increased from patches of 20 WSIs to patches of 621 WSIs, until all model results converged. AUPRC: Area Under the Precision-Recall Curve. (**c**) Performance of various model architectures under the final training set (patches derived from 621 WSIs). The best performance was highlighted in bold. For both NEC and MVP, the best AUC and AUPRC were achieved with the voting results of three models (NEC: AUC of 0.9968 and AUPRC of 0.9788, MVP: AUC of 0.9995 and AUPRC of 0.9860). AUC: Area Under the Receiver Operating Characteristic Curve.

## Materials and Methods

### Data collection

This study was based on data from the Chinese Glioma Genome Atlas (CGGA: 1,968 WSIs) cohort from Beijing Tiantan Hospital and publicly available cohort The Cancer Genome Atlas (TCGA: 1,705 WSIs). The image samples include low-grade glioma (LGG) and high-grade glioma (HGG) which are all digitized hematoxylin and eosin (H&E) stained from formalin-fixed, paraffin-embedded (FFPE) histology slides, referred to Whole-slide images (WSIs). All the image samples with section folds, large areas of pen markings, and poor staining were removed under the review of pathologists. The pathology reports were downloaded from TCGA. The clinical and molecular features of 1,122 TCGA patients were collected from cBioPortal, Mu et al.^3^ and Ceccarelli et al.^22^ (**SMa. 1**).

### WSI and patch preprocessing

Whole-slide images (WSIs) are considerably too large, exceeding 70,000 pixels in each dimension, to be directly used as input for training deep learning models. These circumstances introduce specific complexities within the domain of image analysis. To balance the size of images appropriate for histological patterns detection and training speed, we tiled WSIs into patches with the size of 2,048 X 2,048 pixels with a 50% overlap, resulting in 3,000 to 4,000 patches per WSI, and discarded patches containing small tissue area at 40 X magnification. Meanwhile, for WSIs with a maximum magnification of 20X, the tiled patch size will be set to 1,024 X 1,024 pixels. These patch sizes are appropriate for most WSIs, enabling pathologists to identify crucial diagnostic features. Here we employed the Gaussian smoothing filter and Otsu’s method to differentiate between the tissue area and background area in thumbnail images that compressed from the WSIs, resulting in the generation of binary segmentation masks for each WSI (**Fig. S1a**). A stain normalization algorithm was subsequently applied to the tiled patches to mitigate variations in the staining process. A standard patch was chosen to normalize all other patches to the same staining level (**Fig. S1b**).

### Patch-level datasets amplification and annotation

The annotated dataset is a scarce resource in the field of medical image deep learning. An active learning pipeline was established (**Fig. 2a**) to obtain two datasets for training, validating, and testing the NEC and MVP patch-level models, respectively. In this pipeline, the initial dataset is manually labeled by expert pathologists and consists of 150 positive patches and 150 negative patches. Based on the established dataset, three initial deep learning models were trained and utilized to predict NEC or MVP on a patch pool containing all available patches from the CGGA dataset. With the assistance of pathologists, we were able to randomly verify the predicted results. The incorrectly predicted patches were relabeled and reintegrated into the original dataset. Subsequently, a new model was trained based on the updated dataset. After multiple rounds of active learning, we obtained the NEC dataset comprising 3,552 positive and 8,369 negative patches, as well as the MVP dataset containing 3,139 positive and 11,488 negative patches. Finally, all patches were meticulously reviewed by three pathologists to verify the accuracy of the labels.

### Patch-level and patient-level models

In this study, we used different pre-trained CNN architectures for the necrosis and microvascular proliferation to better accomplish the patch-level classification tasks, including DenseNet201^23^, EfficientNetB2^24^, and ResNet50^25^. The reason for selectin these three typical models is described in detail in **SMe. 1**.

We also proposed a deep learning model, named Probability Localization Network (PLNet) to treat the heatmap as an image to do patient-level classification. The PLNet is a CNN model which is inspired by the skip connections and designed based on the heatmap data characteristics (50% overlap patch tiling) with 3 X 3 kernel size for the first layer feature extraction (**Fig. S4d**).

### Patch-level models training, validation and evaluation

We utilized 80% of patches in the dataset for training and allocated 20% for validation. The networks parameters were initialized with the best set achieved on the ImageNet dataset. The training details of the models were provided in the **SMe. 2**. After completing the training of the classification models, we assessed their performance on an independent dataset. 14 WSIs, which were not involved in data amplification and model training, were tiled into patches and labeled by 3 pathologists. Any patches with disagreements among the pathologists or that lacked consensus on a single patch were excluded. Ultimately, the necrosis independent test dataset comprised 777 positive and 16,941 negative patches, while the microvascular proliferation independent test dataset comprised 485 positive and 14,885 negative patches. All models were evaluated by area under the receiver operating characteristic curve (AUC) scores and area under the precision-recall curve (AUPRC) scores. The Grad-CAM^26^ visualization tool was employed to further verify if the models accurately predicted necrosis and microvascular proliferation based on precise areas. Additionally, for further evaluation, the open-sourced tool QuPath was utilized to visualize the predicted outcomes of all the patches within the WSIs.

### Generating probability localization maps and extracting labels for patient-level NEC and MVP models training and validation

After predicting all the patches within WSIs, we assembled the patch-level predicted results and coordinates to construct the probability localization map for subsequent analysis (**Fig. S4a**). The detailed steps were as follows: First, the positive probabilities, predicted for NEC or MVP, of all patches within a single WSI were compiled. Second, the positive probabilities of all patches were assembled based on their coordinates, which were tiled from the WSI. For instance, in the tiling of a WSI into patches, each patch was considered equivalent to a pixel, thereby converting the WSI into a matrix. Third, because of the assembled matrices with varying sizes for each WSI, which posed challenges in learning the probability distribution map. While a direct approach to address this issue would involve resizing all maps to a uniform size, the resizing process could alter the shape, size, and other characteristics of the NEC or MVP area. To preserve these characteristics, a straightforward method was implemented, matching the matrix to the center of a template with dimensions of 224 X 224 pixels, while setting the blank area to 0 positive probability. The selection of 224 X 224 pixels was based on the statistics of all matrix sizes from CGGA and TCGA, ensuring compatibility with all statuses. Ultimately, the uniform-size probability distribution maps were produced.

To obtain the patient-level labels for WSIs, we proposed a pipeline to employ optical character recognition (OCR) techniques to extract searchable text documents from pathology reports in PDF format downloaded from TCGA website. Additionally, we incorporated a fuzzy keyword search for necrosis and/or microvascular proliferation related diagnoses to acquire patient-level labels for the WSIs (**Fig. S4b**). For NEC, 95 positive and 156 negative patients, and for MVP, 100 positive and 145 negative patients were included in this processing. The probability localization map integrated previously was used to train and valiate our proposed PLNet (Probability Localization Network) using 5-fold cross-validation (**SMe. 3**).

### NEC and MVP pattern clustering with differential expression and gene set enrichment analyses

Necrosis and microvascular proliferation are two of the most potent predictors of poor prognosis in adult diffuse gliomas. Beyond their roles as mere morphological markers, these structures are undeniably pivotal in driving the accelerated growth properties that define the transition from low grade to high grade. To further explore the characteristics of NEC and MVP, as well as patients with NEC and/or MVP, the mini-batch K-Means method was employed to cluster 11,923 NEC positive patches and 19,297 MVP positive patches tiled from 1,699 TCGA WSIs. One patch was represented by the combined 2,272-dimensional feature vectors extracted from the 3 CNN patch-level models previously trained (DenseNet201: 896, EfficientNetB4: 352, ResNet50: 1,024). Principle component analysis (PCA) was subsequently employed to reduce the feature vector dimension to 100 for subsequent clustering. All these patches were grouped into 2 clusters to differentiate between pseudopalisading necrosis (PAN) enriched patients and other necrosis enriched patients or between glomeruloid microvascular proliferation (GMP) enriched patients and other MVP enriched patients using the Mini-Batch K-Means method. Additionally, UMAP^27^ was utilized for visualizing the clustering outcomes. All patients were categorized into groups based on the enrichment of specific patches within their WSIs (**SMe. 4**).

We conducted differential expression with volcano plot and gene set enrichment analysis (GSEA)^28, 29^ with the normalized RNA-seq expression data acquired for the TCGA glioma dataset, which were selected samples from two groups based on the PAN clustering result: PAN enriched patients (n=42) and other necrosis enriched patients (n=70), and GMP clustering result: GMP enriched patients (n=56) and other MVP enriched patients (n=71) (Details in **SMe. 5**). Only Hallmark gene sets^30, 31^ with a p-value < 0.05 were selected as the functionally enriched biological states or processes.

## Results

### Patch-level models classification performance

Commencing with a modest dataset comprising 838 patches from 20 WSIs, we separately trained 3 distinct common CNN architectures, which were DenseNet201^23^, EfficientNetB2^24^, and ResNet50^25^, for the NEC and MVP patch-level classification tasks. Subsequently, after updating the training dataset with models and pathologists, we retrained the 3 CNN models. AUC and AUPRC of all models were evaluated on the independent test dataset, comprising 17,718 NEC and 15,370 MVP patches from 14 WSIs that were excluded from training dataset. The annotations of all patches in the training and test datasets were independently reviewed by three neuropathologists (**Fig. 2a**). We evaluated the improvement in model performance as the training dataset size increased during active learning (**Fig. 2b**). The voting results represented the median value of the 3 models from each dataset. The results indicated that model performance improved with the inclusion of more WSIs in the training dataset, ultimately reaching the limitation of label annotation accuracy. Finally, two well-annotated datasets were established for training the NEC and MVP patch-level models. These datasets include 3,552 positive patches and 8,369 negative patches for NEC, and 3,139 positive patches and 11,488 negative patches for MVP. AUC and AUPRC results indicated that models exhibited high performance (Table in **Fig. 2c**), notably achieving an AUC of 0.9968 and AUPRC of 0.9788 for NEC models’ voting results, and an AUC of 0.9995 and AUPRC of 0.9860 for MVP models’ voting results.

### Quantitative analysis in WSIs and visual explanations in patches

Previously, conducting quantitative analysis of histological features was excessively time-consuming for pathologists during diagnosis. To tackle this issue, we shifted our focus to whole-slide images and investigated whether CNN models could be leveraged to quantitatively analyze histological features from WSIs, like the percentage of NEC and MVP positive patches in one WSI. For this purpose, we selected a WSI from the independent test set to demonstrate the quality of quantitative analysis of NEC and MVP compared to the annotation made by pathologists, as illustrated in **Fig. 3a** and **Fig. S2b, c**. To evaluate the predicted performance of the CNN models, we tasked pathologists with independently classifying all patches from the WSIs through visual inspection. The results revealed that the CNN model could accurately detect and quantify NEC and MVP with predicted performance compared to pathologists. The overall NEC and MVP positive patches identified by the CNN models highly overlaps with the positive patches labeled by pathologists (voting results: **Fig. S2a,** separate results: **Fig. S3**). In this WSI, the precisions for NEC and MVP patches were 1 and 0.9726, respectively, while the sensitivities were 0.9375 and 0.9342. Furthermore, the model accurately calculated the percentage of NEC and MVP positive patches, demonstrating results comparable to those of the pathologists as a quantification metric for patient diagnosis.

**Figure 3.**
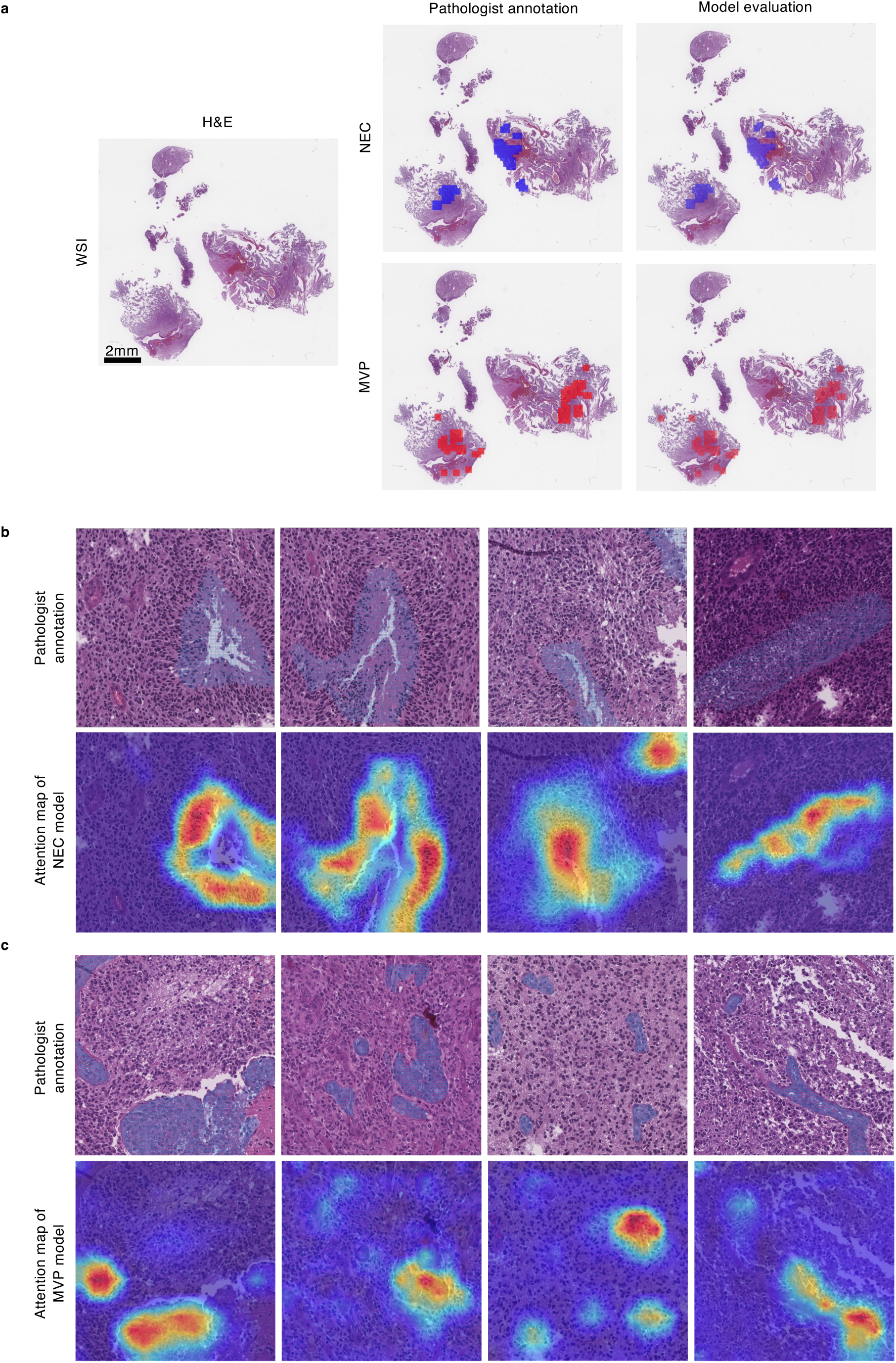
Performance of the patch-level deep learning models. (**a**) A WSI in the independent test set was reviewed to demonstrated the model performance (left panel). In the middle panel, the dark blue (or red) areas highlighted the NEC (or MVP)-positive patches classified by pathologists through visual inspection. The right panel included the results of the voting models with light blue (or red) areas highlighting the NEC (or MVP)-positive patches by the deep learning models. (**b**-**c**) Grad-CAM analysis of key pathology features. NEC (**b**) and MVP (**c**) were manually highlighted by pathologists (blue area in the top panels), while Grad-CAM analyses of the deep learning model provided the class-discriminative visualization of the corresponding features shown in the heatmap in the bottom panels.

Given the satisfactory predicted performance of the classification models, it is essential to delve deeper into how the deep learning model arrived at its predictions and ascertain whether the model genuinely learned the meaningful histopathological features. To address these questions, we carried out gradient-weighted class activation map analysis (Grad-CAM) on the CNN models. 4 NEC and 4 MVP positive patches were randomly selected for these assessments. As depicted in **Fig. 3b**, **c**, the localization of specific features that CNN models focused on during prediction aligned well with the localization of NEC and MVP identified by the pathologists. These alignments indicate that the CNN models trained on our datasets successfully learned meaningful histological features and utilized the information for prediction.

### Patient-level models for computer-aided diagnosis

We conducted analyses utilizing the generated probability localization maps and extracted labels for patient level. The caption content of all the pathology reports generated by our pipeline is illustrated in **Fig. S4c**. Each patch was assigned a predicted probability for NEC or MVP and positioned based on its coordinates within the WSI. Subsequently, a heatmap of probability localization would be generated. We proposed a deep learning model, named PLNet (Probability Localization Network, **Fig. S4d**), to treat the heatmap as an image for patient-level classification. The datasets contained gliomas ranging from WHO grade II to grade IV in TCGA cohort (Table in **Fig. 4a**). The patient-level predicted labels were directly derived from the patch-level outcomes, where the labels were determined based on whether the whole-slide image included predicted positive NEC or MVP patches. Utilizing pathology reports as the ground truth, accuracy (NEC: 81.27%, MVP: 87.76%) and sensitivity (NEC: 55.79%, MVP: 70%) were calculated for TCGA WSIs (**Fig. 4b**). The pipeline for the patient-level aggregation models is illustrated in **Fig. 4c**. Accuracy and sensitivity were determined by setting different probability thresholds for TCGA WSIs, using pathology reports as ground truth. Additionally, they were evaluated through 5-fold cross-validation across all the data. The results demonstrated that our PLNet model achieved the highest accuracy of 88.05% for NEC and 90.20% for MVP, along with a sensitivity of 73.68% for NEC and 77% for MVP. When the sensitivity was set at 80%, the accuracy for NEC reached 79.28% and for MVP reached 77.55% (**Fig. 4d**). Compared to **Fig. 4b**, both accuracy and sensitivity were enhanced for NEC and MVP. The results from TCGA datasets also indicated that our deep learning model, trained on CGGA datasets, possessed the capability to generalize effectively in identifying and quantifying NEC and MVP across independent cohorts.

**Figure 4.**
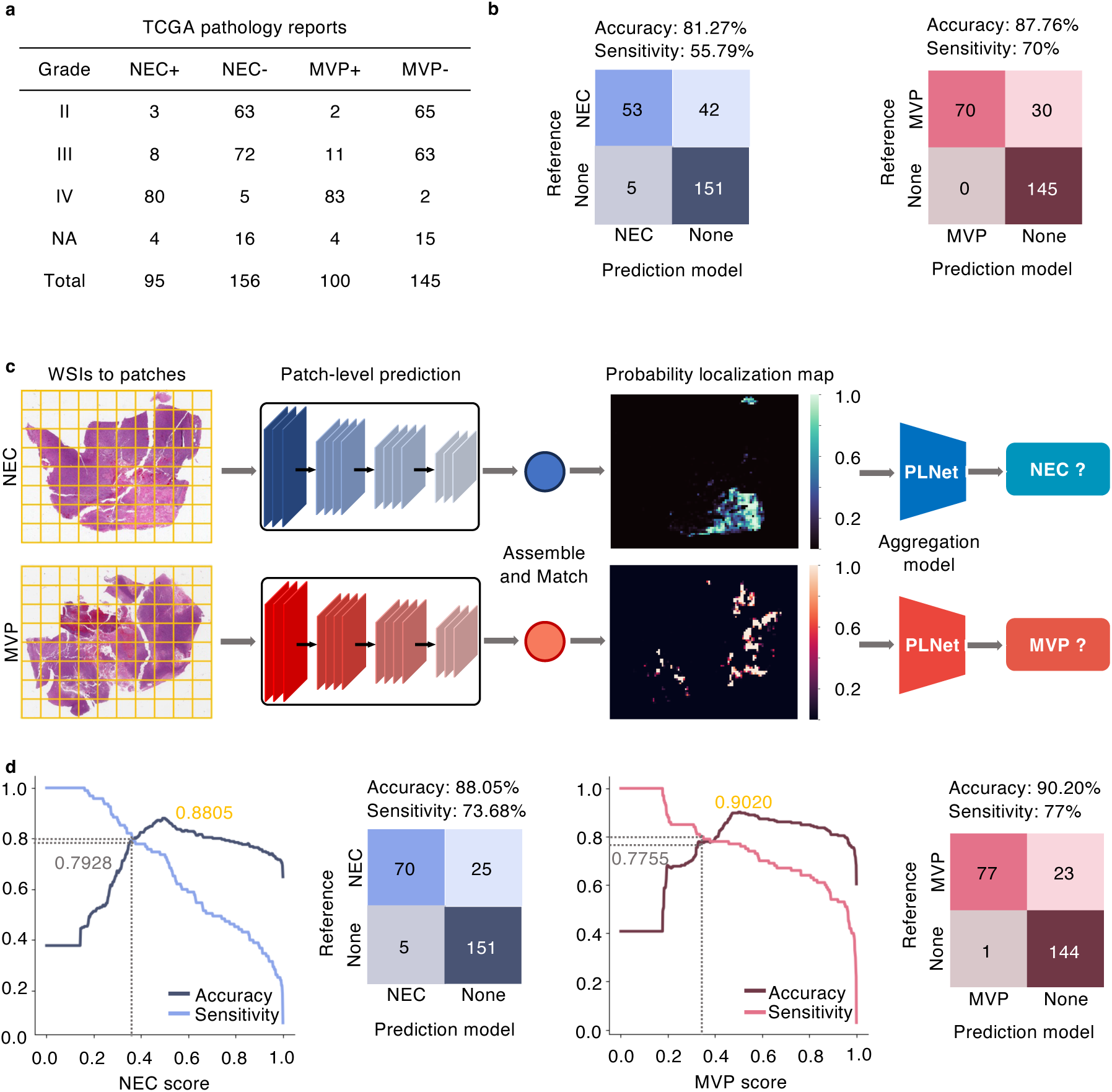
Patient-level models of NEC and MVP. (**a**) NEC and MVP annotations extracted from TCGA pathology reports. (**b**) Accuracy and sensitivity of patch-level models to predict TCGA pathology reports. The patient-level prediction label derived from the patch-level results, whether the WSI has positive NEC or MVP patches as the patient-level label. Using pathology reports as ground truth, accuracy and sensitivity were respectively calculated for TCGA WSIs (NEC: accuracy of 81.27% and sensitivity of 55.79%, MVP: accuracy of 87.76% and sensitivity of 70%). (**c**) Pipeline for the patient-level aggregation models. First, we utilized our trained patch-level models to predict the tiled patches form TCGA WSIs. Each patch was assigned a predicted probability for NEC or MVP and positioned according to its coordinates within the WSI. Subsequently, a probability localization map would be generated. We then proposed a deep learning model called PLNet (Probability Localization Network) to treat the map as an image for patient-level classification. (**d**) NEC and MVP AUC results of 5-fold cross-validation for TCGA cohort of patients based on our proposed PLNet. The results demonstrated that our PLNet model achieved the highest accuracy of 88.05% for NEC and 90.20% for MVP, along with a sensitivity of 73.68% for NEC and 77% for MVP. When the sensitivity was set at 80%, the accuracy for NEC reached 79.28% and for MVP reached 77.55%. Compared to (**b**), both accuracy and sensitivity were enhanced for NEC and MVP.

### Survival analysis for outcomes of patient-level models

We further investigated whether our patient-level model could be utilized for prognosis prediction. Following the diagnosis guideline in conjunction with the NEC and MVP predicted results and pathology reports, these 878 patients with WSIs from 1,122 patients were categorized into four groups: Oligodendroglioma (IDH-mutant and 1p/19q-codeleted, WHO Grade 2/3), Astrocytoma (IDH-mutant, 1p/19q-non-codeleted and CDKN2A/B wildtype, WHO Grade 2/3), Astrocytoma (IDH-mutant, 1p/19q-non-codeleted and CDKN2A/B HD, WHO Grade 4), Glioblastoma (IDH wild type, WHO Grade 4). Using our patient-level model, we conducted several analyses to verify its practicality and clinical potential. We performed a correlation analysis between the true labels from pathology reports and the model predicted scores (**Fig. 5a, b**). For NEC, when the model predicted score was less than or equal to 0.5 (n=173), the survival probabilities of patients were similar (*P*=0.161) to those with a true label of 0 (n=153). When the model predicted score was greater than 0.5 (n=74), the survival probabilities of patients were similar (*P*=0.818) to those with a true label of 1 (n=94). For MVP, when the model predicted score was less than or equal to 0.5 (n=165), the survival probabilities of patients were similar (*P*=0.097) to those with a true label of 0 (n=142). When the model predicted score was greater than 0.5 (n=77), the survival probabilities of patients were similar (*P*=0.772) to those with a true label of 1 (n=100). Additionally, our model could effectively classify patients into two groups with distinct survival predictions (*P*=1.1596E-13 for NEC and *P*=3.7353E-12 for MVP between predicted scores ≤ 0.5 and > 0.5). Subsequently, we utilized various model predicted scores as thresholds to further divide the samples into two groups: one group with scores less than or equal to the threshold and the other group with scores greater than the threshold (**Fig. 5c**). We calculated the survival differences between these two groups and determined the corresponding p-values. For NEC samples with a true label of 0, we identified four thresholds (0.34, 0.35, 0.36, 0.37) where their -log10 p-values were greater than -log10 0.05. The highest -log10 p-value was observed at 0.34, where 22 samples with model-predicted scores less than or equal to 0.34 had better survival (**Fig. 5d**, *P*=0.031, -log10 *P*=1.508) compared to 131 samples with scores greater than 0.34. We further extended our analysis to include all TCGA data with WSI and observed that our model can effectively differentiate between survival outcomes (**Fig. 5e, g**). For type A, IDHmut-non-codel, when our model predicted value is less than or equal to 0.66 or greater than 0.66 for NEC, or when our model predicted value is less than or equal to 0.33 or greater than 0.3 for MVP, the group with lower values exhibits superior survival compared to the group with higher values (**Fig. 5f, h**, *P*=0.001 for NEC and *P*<0.001 for MVP). Based on this, for A, IDHmut-non-codel CDKN2A/B wildtype records, we further subtyped the samples with more significant NEC and/or MVP characteristics. These records, derived from 186 CDKN2A/B wildtype samples across all A, IDHmut-non-codel samples in TCGA, have the potential to be a higher grade (**Fig. S5b**). Similar results were also observed in type O, IDHmut-codel (**Fig. S5a, c**). We updated the Grade (WHO 2021) based on our model as shown in Supplementary Materials.

**Figure 5.**
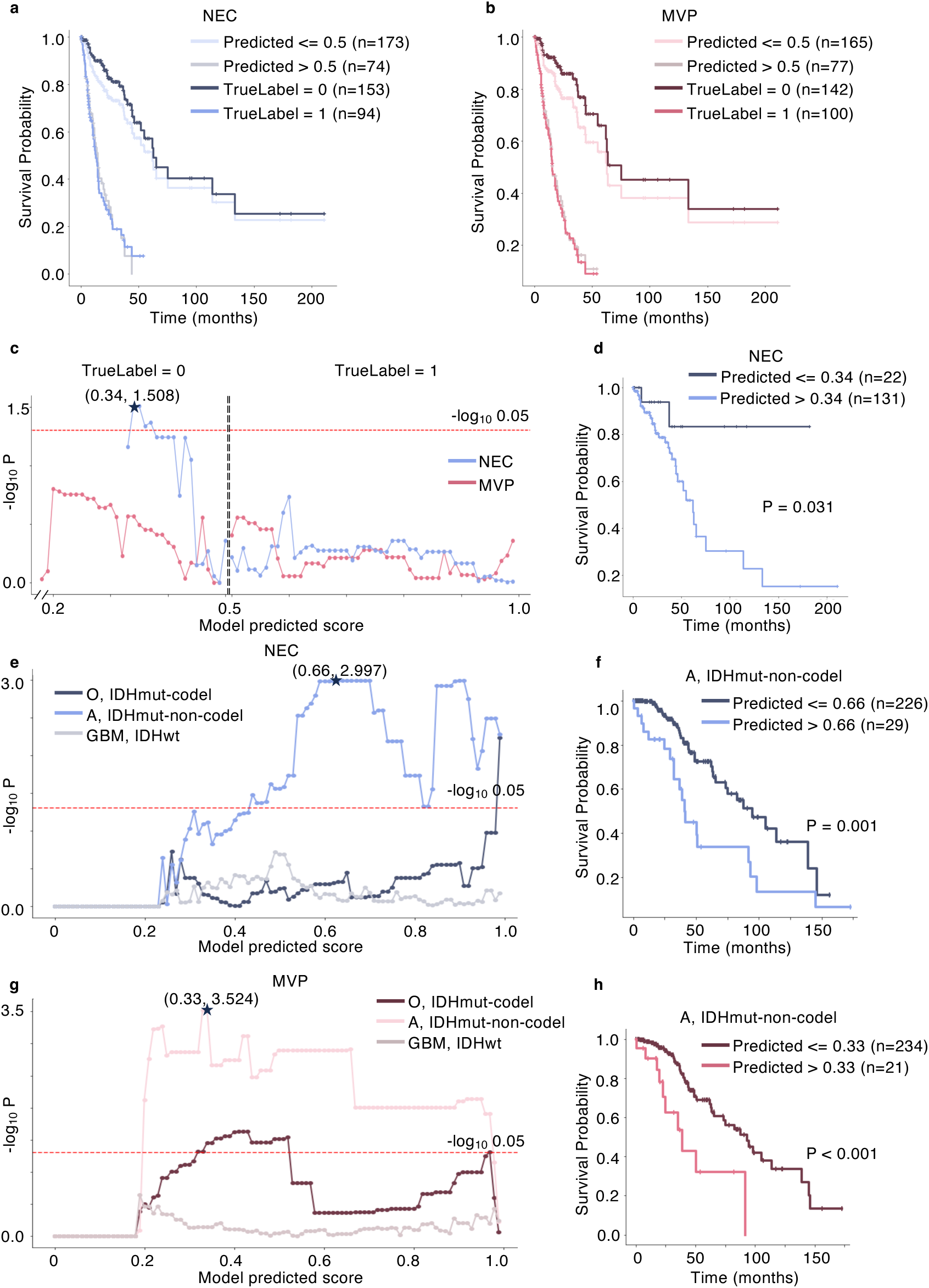
Survival analysis using the patient-level model. (**a**)-(**d**) are based on data with reports, while (**e**)-(**h**) are based on all TCGA data. (**a**) Survival difference analysis for NEC between the true label and the model-predicted score. (**b**) Survival difference analysis for MVP between the true label and the model-predicted score. (**c**) By using different model-predicted scores as thresholds, we divide the samples into two groups. The survival difference between these two groups is calculated, and the corresponding p-value is determined. (**d**) Survival curves based on the most significant p-value from (**c**). This demonstrates that our model-predicted scores can further subtype the true labels. (**e**) In different molecular subtypes, the p-values of survival curves using different probability prediction thresholds of the NEC model. O: Oligodendroglioma, A: Astrocytoma, GBM: Glioblastoma. (**f**) Survival curves based on the most significant p-value for A, IDHmut-non-codel from (**e**). (**g**) In different molecular subtypes, the p-values of survival curves using different probability prediction thresholds of the MVP model. (**h**) Survival curves based on the most significant p-value for A, IDHmut-non-codel from (**g**).

### Clustering-based analyses with feature extraction

Uniform Manifold Approximation and Projection (UMAP)^27^ demonstrated pseudopalisading necrosis (PAN) patches enriched in one cluster of all NEC patches, and glomeruloid microvascular proliferation (GMP) patches also enriched in one cluster of all MVP patches, which were manually checked by pathologists (**Fig. 6a**). Patients enriched with PAN or GMP were differentiated from all TCGA patients based on the quantity of PAN or GMP patches present in each WSI. Kaplan-Meier analysis was utilized to assess whether the PAN and GMP enriched patients could be effectively discriminate from other NEC and MVP enriched patients, respectively. The results revealed that the PAN (n=128) and GMP (n=178) enriched patients exhibited poorer survival outcomes compared to other NEC (n=120) and MVP (n=135) enriched patients, respectively, with p-values of 1.04E-04 for the NEC group and 1.30E-03 for the MVP group (**Fig. 6b**). We extended our analyses by conducting differential expression analysis with volcano plot (**Fig. 6c**), and gene set enrichment analysis (GSEA) (**Fig. S6a-c**) for both the PAN enriched patients compared with the other NEC enriched patients, as well as for the GMP enriched patients compared with the other MVP enriched patients. In the volcano plots (**Fig. 6c**) comparing the PAN enriched patients with other NEC enriched patients, several genes exhibit significant upregulation in the PAN enriched patient group, including FEZF1, SEPTIN14, SCNN1B, and IRX5. Conversely, notable downregulated genes such as PCDHGA11, KRT14, CCL19, and NMUR2 are observed in the PAN enriched patient group. Comparing the GMP enriched patients with other MVP enriched patients, several genes exhibit significant upregulation in the GMP enriched patient group, including CDK4, TSPAN31, CYP27B1, and PRDM13. Conversely, notable downregulated genes such as SELL, KRT6A, SMCP, and CCL21 are observed in the GMP enriched patient group. These upregulated genes suggest potential pathways or mechanisms that are more active in the PAN or GMB enriched patient group compared to the other NEC or MVP enriched patient group. The downregulation of these genes may indicate suppressed pathways or functions in the PAN or GMB enriched patient group, potentially highlighting areas of biological significance or therapeutic interest. The gene set enrichment analysis (GSEA) based on Hallmark gene sets revealed the significantly altered pathways (**Fig. S6a, b**). For p-value less than 0.05, the 7 enriched pathways in the PAN enriched patient group are primarily associated with cellular proliferation related, including E2F targets, G2M checkpoint, MYC targets V1, and Mitotic spindle, metabolic related like Glycolysis, pathway related like Hypoxia, and signaling related like MTORC1 signaling (**Fig. S6a**). The enrichment of these pathways in the PAN enriched patient group suggests potential alterations in cell cycle regulation, metabolism, cellular response to oxygen levels, and signaling pathways related to growth and proliferation. This molecular profile may indicate specific biological processes, cellular adaptations, or dysregulations that are characteristic of this group being studied. Meanwhile, the 16 enriched pathways in the GMP enriched patient group are primarily associated with cellular proliferation related, including E2F targets, G2M checkpoint, MYC targets V1, V2, and Mitotic spindle, metabolic related like Glycolysis, and pathway related such as Hypoxia and Unfolded protein response, development related such as Angiogenesis, Epithelial mesenchymal transition, Spermatogenesis, and Pancreas beta cell, DNA damage related like DNA repair, immune related like Interferon alpha response, and signaling related such as MTORC1 signaling, and TNFA signaling via NFKB (**Fig. S6b**). The presence of these pathways in the GMP enriched patient group suggests a complex interplay of cellular processes related to cell cycle regulation, metabolism, stress responses, angiogenesis, epithelial-mesenchymal transition, specialized cell functions, DNA integrity maintenance, immune responses, and signaling pathways. This intricate network of pathways may collectively contribute to the unique molecular profile and potentially the underlying biology of this group being studied. Previous studies investigated that PAN in glioblastoma was associated with hypoxia and GMP in glioblastoma was associated with angiogenesis^32, 33, 34, 35, 36^, which were consistent with our findings. The GSEA results also indicated that the highest normalized enrichment scores were observed in the E2F targets pathway for both PAN and GMP enriched patients (**Fig. S6c**). Furthermore, we conducted more detailed analyses and obtained more sufficient information. Ultimately, we combined all the NEC and MVP predicted results together for generating diagnosis reports (**Fig. S7a**). Additionally, we utilized the patch-level predictions to generate the patient-level heatmap for online demonstration (**Fig. S7b**).

**Figure 6.**
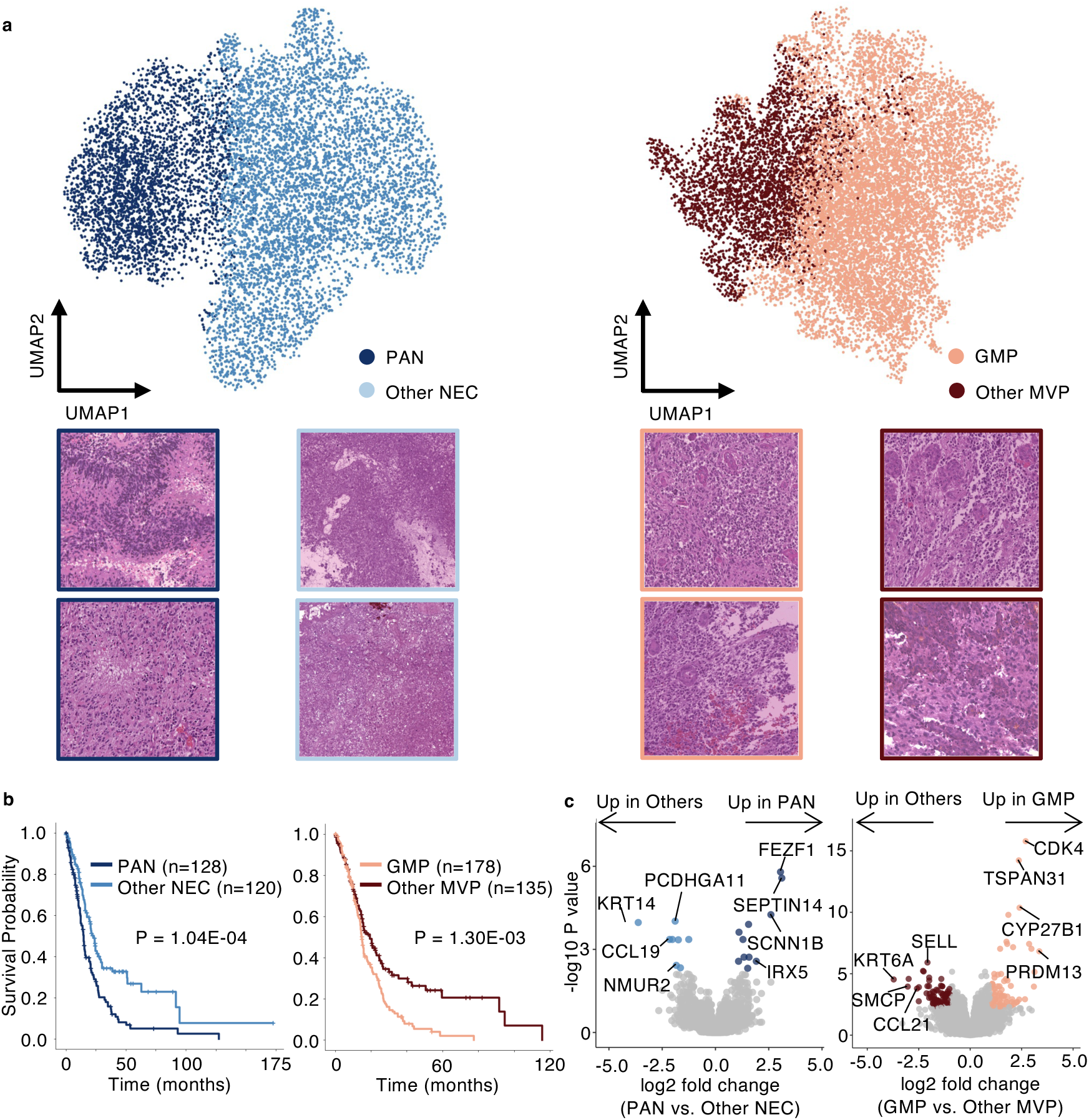
Clustering of NEC and MVP patches and their subtype analysis. (**a**) UMAP projection of K-Means clustering of all 11,923 NEC and 19,297 MVP concatenated 2,272-dimensional feature vectors (DenseNet201: 896, EfficientNetB4: 352, ResNet50: 1,024) of positive patches of TCGA WSI dataset. 2 typical PAN (pseudopalisading necrosis) patches, and 2 other NEC patches were evaluated by pathologists. PAN patches were enriched in dark-blue cluster, and other NEC patches were enriched in light-blue cluster. 2 typical GMP (glomeruloid microvascular proliferation), and 2 other MVP patches were evaluated by pathologists. Other MVP patches were enriched in dark-red cluster, and GMP patches were enriched in light-red cluster. (**b**) Survival analysis for PAN (n=128) vs. Other NEC (n=120) and GMP (n=178) vs. Other MVP (n=135) enriched patients. Both NEC and MVP groups demonstrated significant survival differences. (**c**) Volcano plots depicting differentially expressed genes in PAN and GMP enriched patients.

## Discussion

In this study, we proposed an active learning pipeline to annotate patch-level datasets and train patch-level models for classifying and quantifying NEC and MVP. Subsequently, we proposed a patient-level CNN classification model named PLNet (Probability Localization Network), which was integrated with 3 patch-level CNN models to identify and quantify tumor NEC and MVP from histopathological glioma patients. The high predicted performance of the patch-level and patient-level models indicate that AI-based automatic histopathological diagnosis has the potential to be applied in clinical practice, achieving performance comparable to that of pathologists. The models can improve the efficiency and consistency of the diagnosis process by assisting pathologists in repetitive detection tasks within whole-slide images (WSIs). Moreover, they have the potential to adjust previously uncertain patient grades to updated grades, thereby facilitating more precise diagnoses. We further clustered NEC and MVP positive patches into different subtypes using high-level features extracted by patch-level CNN models to characterize patients enriched with PAN and other NEC, and with GMP and other MVP. The results suggest that deeper information correlated with tumor progression stage and patients’ genomic features can be dug out by our model. This information can be ignored in clinical diagnosis and is difficult to obtain through traditional pathology methods. It would be a starting point to develop an AI-based diagnosis and treatment suggestion system that combines digital pathology and genomics, which can really provide precision medicine specific to each patient.

We have evaluated our models on different datasets and a large number of patients to enhance their generalization ability. However, the diversity of WSIs introduced by variations in tissue collection methods, fixation technologies, levels of hematoxylin and eosin (H&E) staining, different digital slide scanners, various data sources and collecting time makes it challenging to apply the models to every dataset. Our approach demonstrates a promising pipeline from dataset construction, training, evaluating, for further analysis on WSIs. Hopefully, it is feasible to adapt this pipeline to many other diseases.

We utilized formalin-fixed, paraffin-embedded (FFPE) WSIs in this study as they are commonly preferred in pathology diagnosis practices. Another type of WSI is based on frozen tissue, which can be rapidly produced during surgery to aid the surgeon in determination whether the tumor margins are clean. Most of the frozen tissue WSIs will exhibit tissue damaged, displaying a Swiss-cheese-like appearance, but can still provide pathological features for diagnosis. Several previous studies have explored using both frozen tissue and FFPE WSIs as inputs for deep learning models in tumor diagnosis^12, 37, 38^.

Necrosis and microvascular proliferation (MVP) are two primary pathological features in glioma diagnosis. Additional features such as mitosis activity^39, 40, 41^, nuclei atypia, and nuclei density^42, 43, 44^ are also crucial for a comprehensive diagnosis. We are utilizing the approach presented in this paper for the automatic detection and quantification of all these features. With all features automatically detected by the models, we can integrate them to generate a comprehensive diagnostic report that includes all quantified features, genomic indications, treatment suggestions, and potential drug choices. This holds promise for the future of precision medicine era.

## Supporting information

Supplemental Mauscript, Figures and Table

## Data Availability

All data produced in the present study are available upon reasonable request to the authors

## Acknowledgments

The work described in this paper was supported by a grant from the Research Grants Council of the Hong Kong Special Administrative Region, China (Project No. R6003-22). This work was also partially supported by ITC grant (MHP/004/19, ITCPD/17-9) and a seed fund of the Big Data for Bio-Intelligence Laboratory (Z0428) from The Hong Kong University of Science and Technology. J. Wang is also supported by Padma Harilela Professorship.

## Author Contributions

J. Wang and T. Jiang conceived the project and designed the experiments. X. Liu and T. Jiang obtained the patient samples and scanned the WSIs. W. Zou, X. Liu and F. Qiu labeled the necrosis and microvascular proliferation dataset. Y. Guo and H. Huang performed the model training and testing and the dataset correction. Y. Liu and Y. Guo perform survival analysis. Y. Guo performed the differential expression, and gene set enrichment analyses. Y. Guo, H. Huang, Y. Liu and J.W. wrote the manuscript with input from all authors.

## Data sharing

CGGA data, including WSIs (not yet), tumor types and demographics, are available at http://www.cgga.org.cn/. Registration and acceptance of terms and data use agreements are required for access. The human LGG and GBM diagnostic slide images were derived from the TCGA Research Network at https://portal.gdc.cancer.gov/. The code is available for academic research purposes via GitHub at https://github.com/WangLabHKUST/NMADG.

