## Supplemental Mauscript, Figures and Table for "Deep learning-based identification of necrosis and microvascular proliferation in adult diffuse gliomas from whole-slide images": Nec_MVP_Supplementary_Figures.pdf

#### Supplementary figure 1 (related to figure 2)

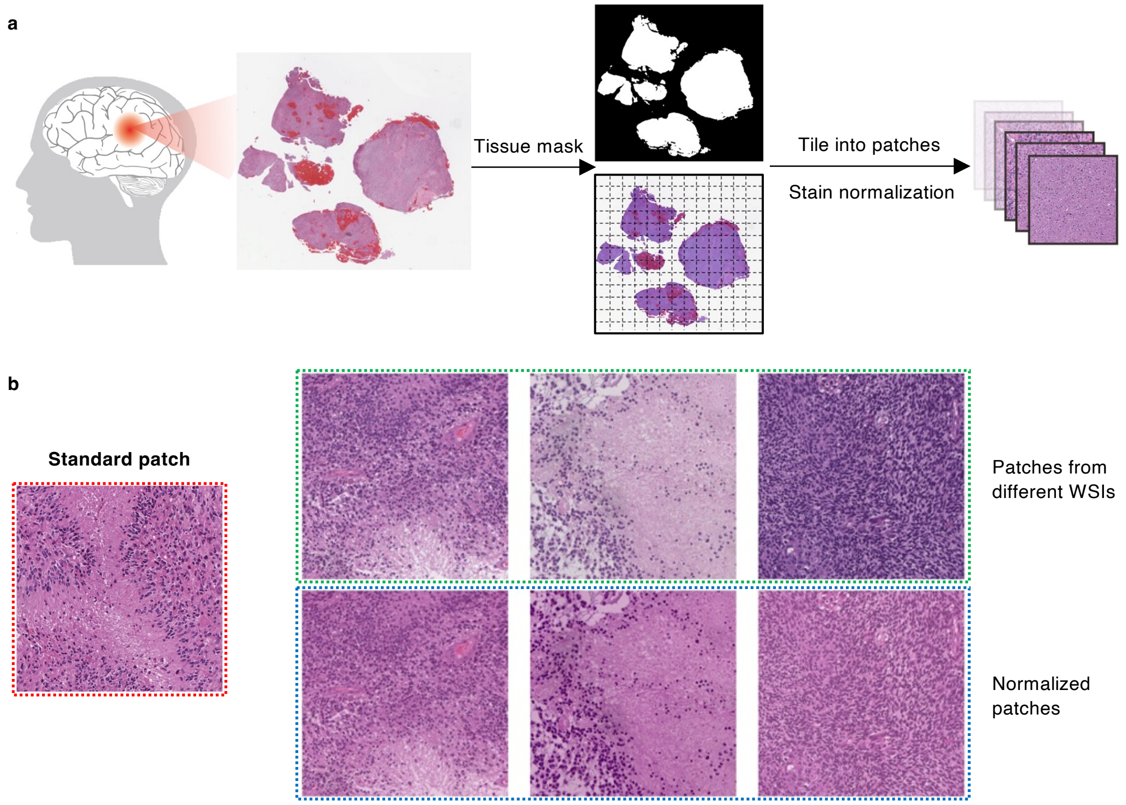

#### Supplementary figure 2 (related to figure 3)

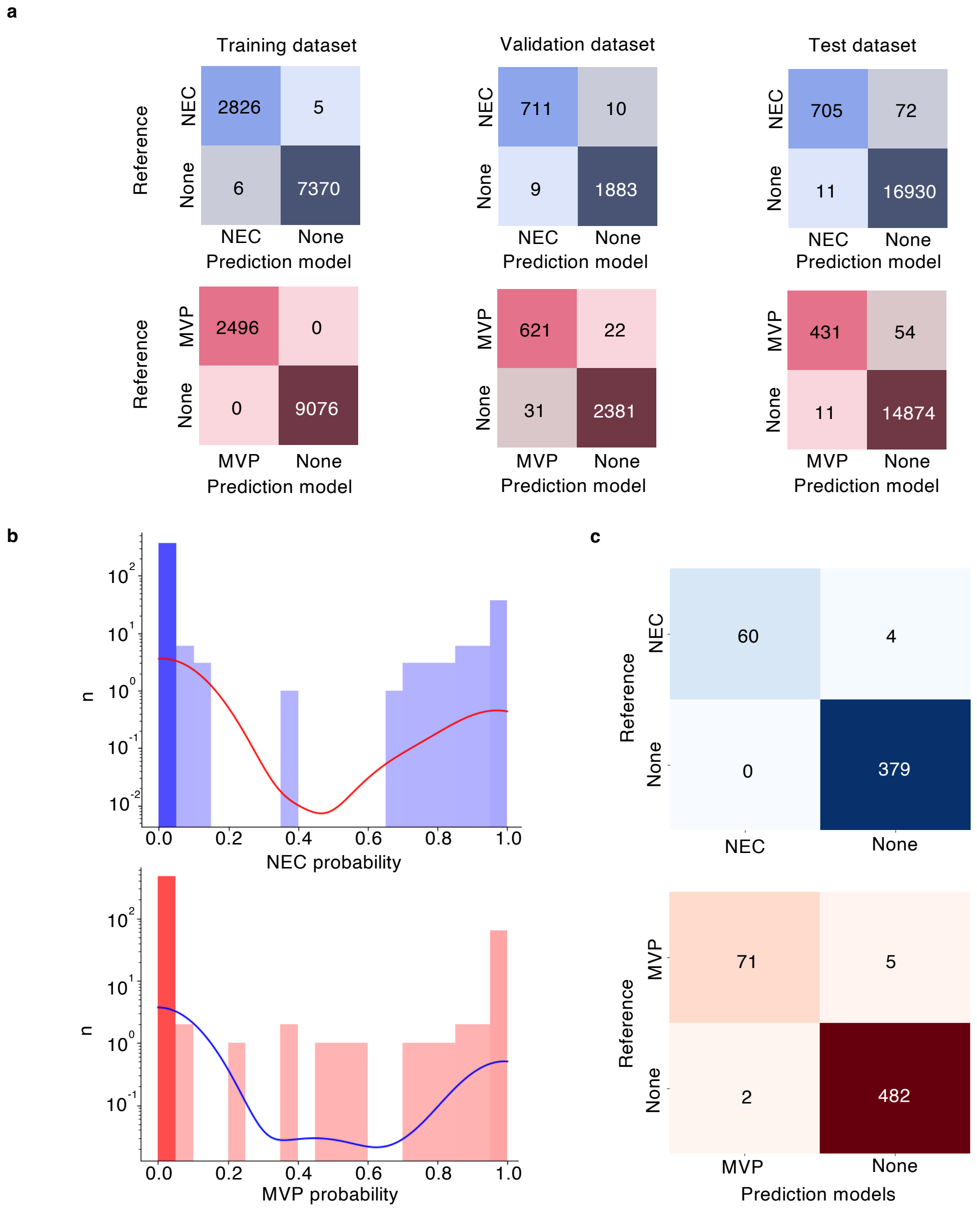

#### Supplementary figure 3 (related to figure 3)

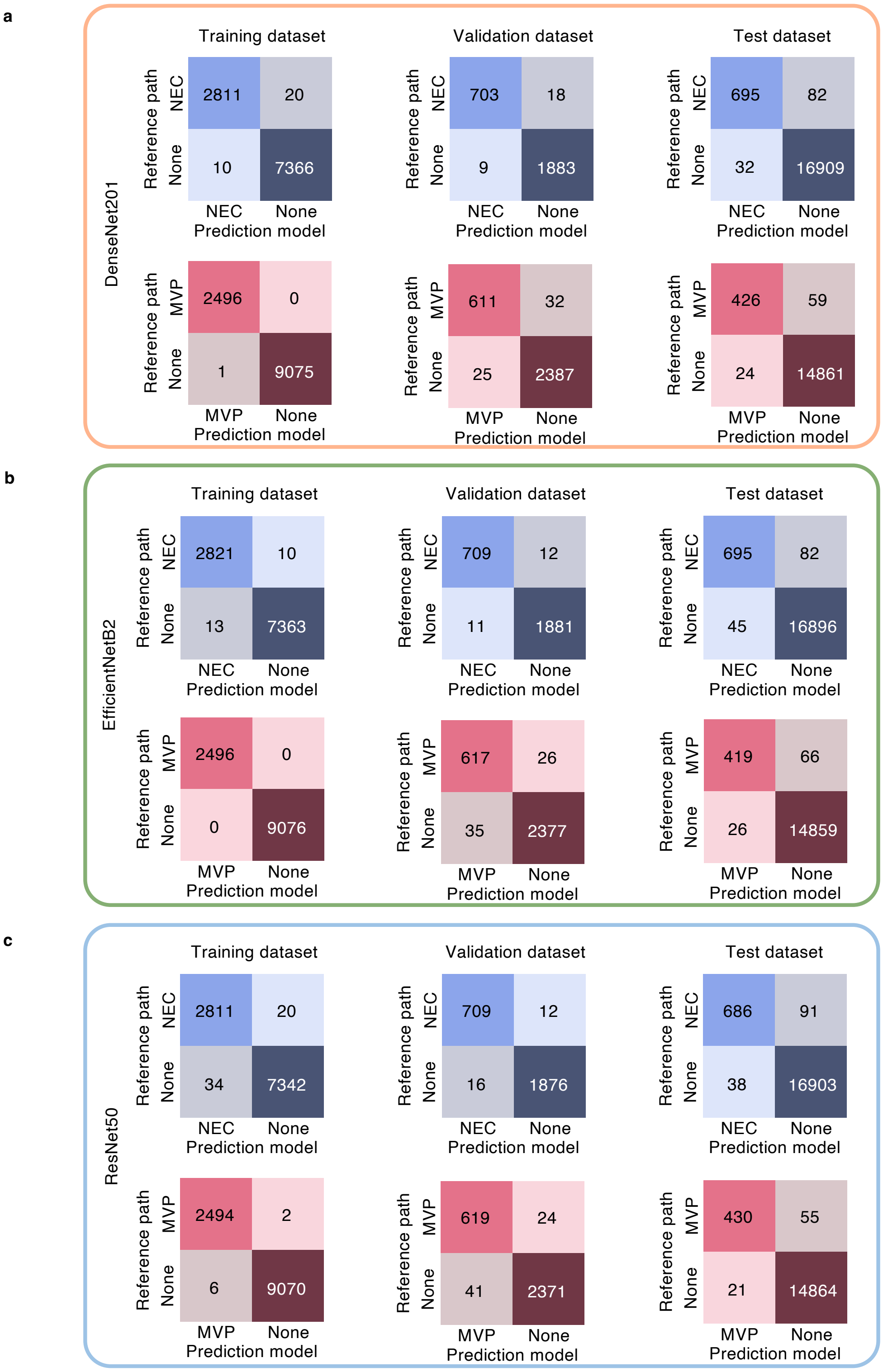

#### Supplementary figure 4 (related to figure 4)

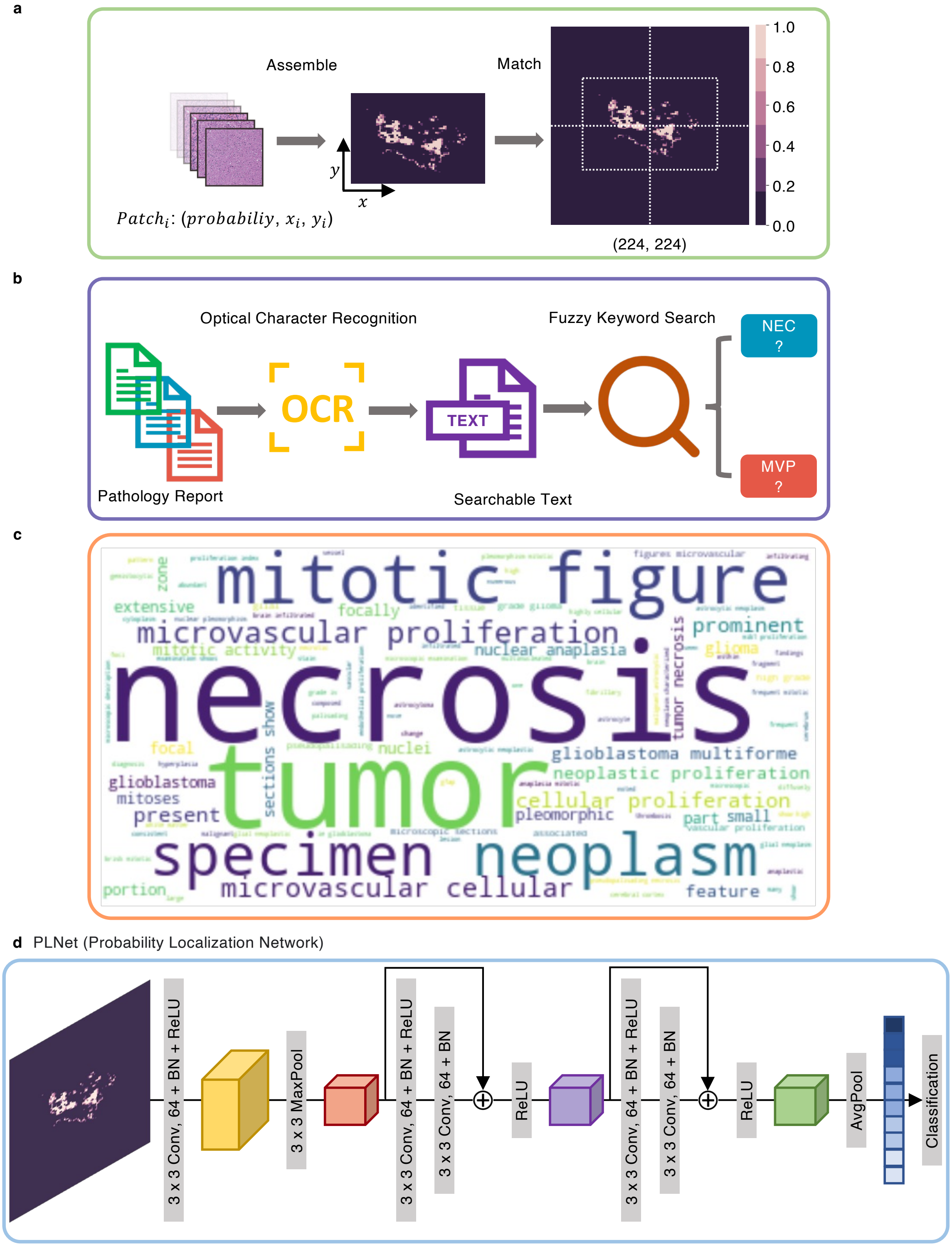

### Supplementary figure 5 (related to figure 5)

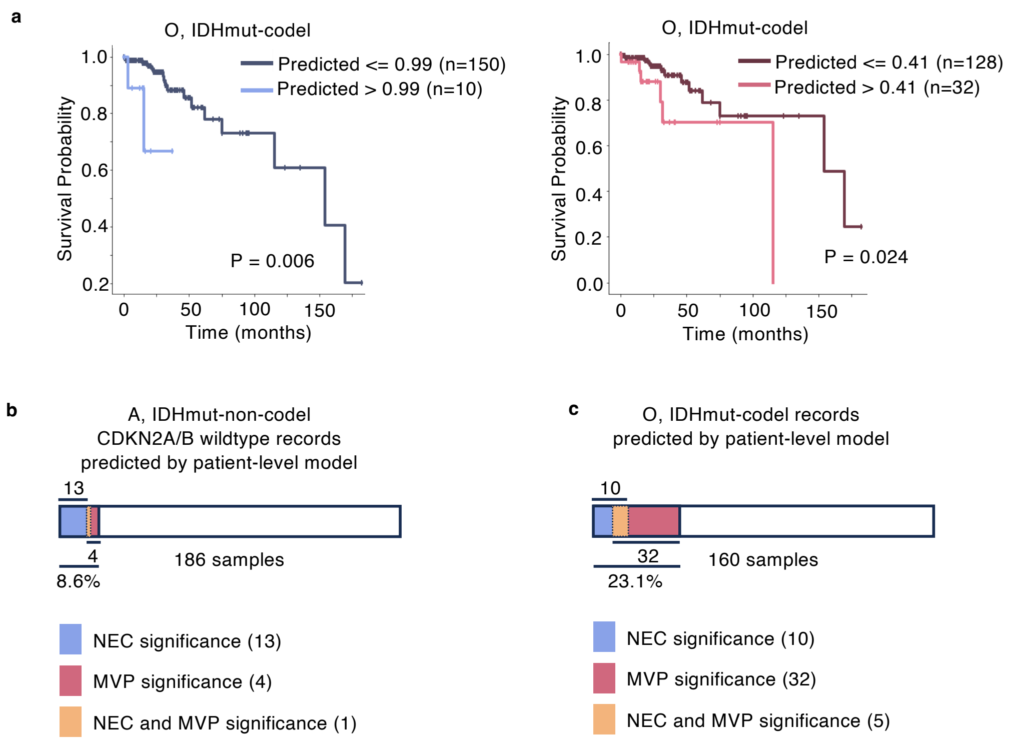

#### Supplementary figure 6 (related to figure 6)

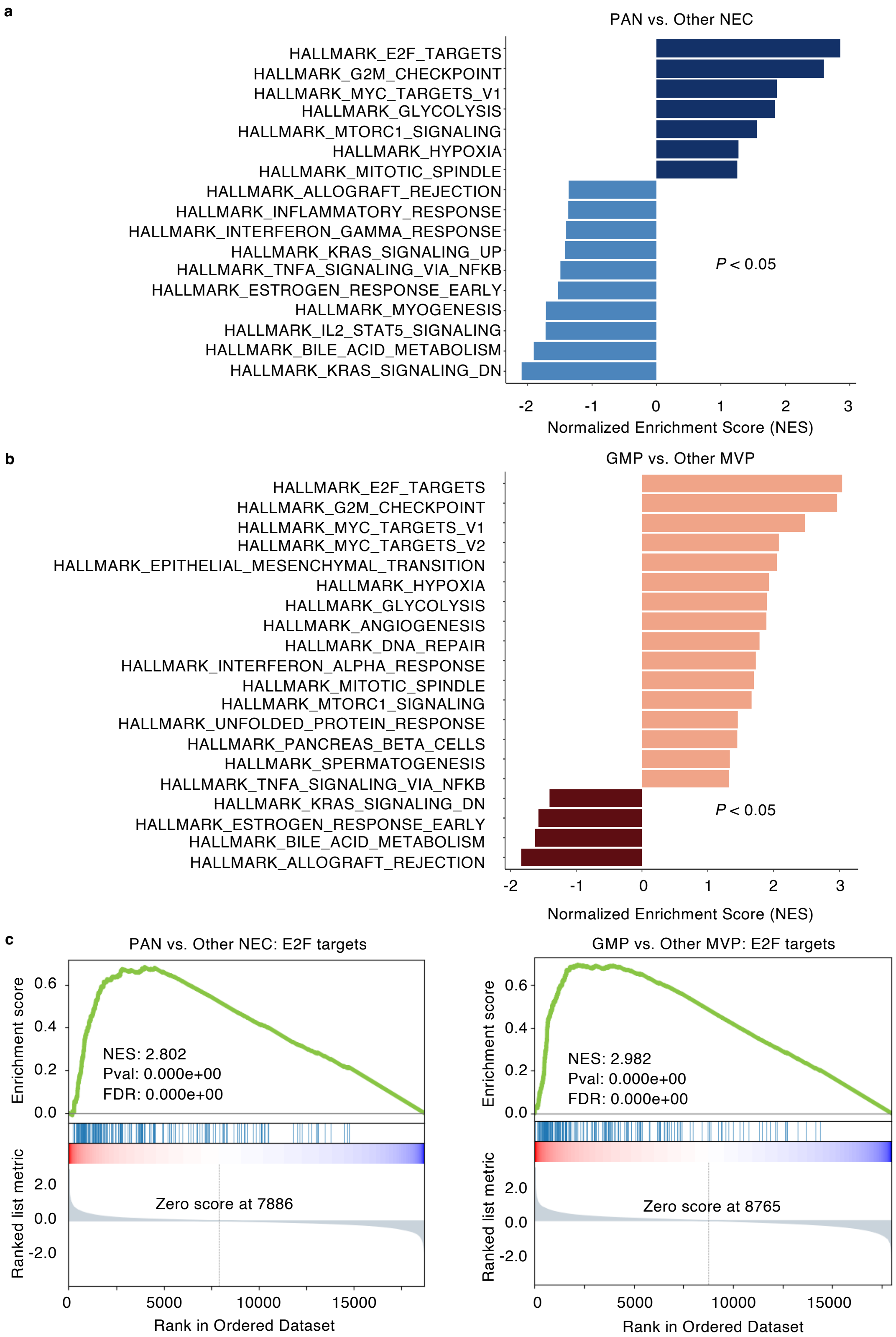

#### Supplementary figure 6 (related to figure 6)

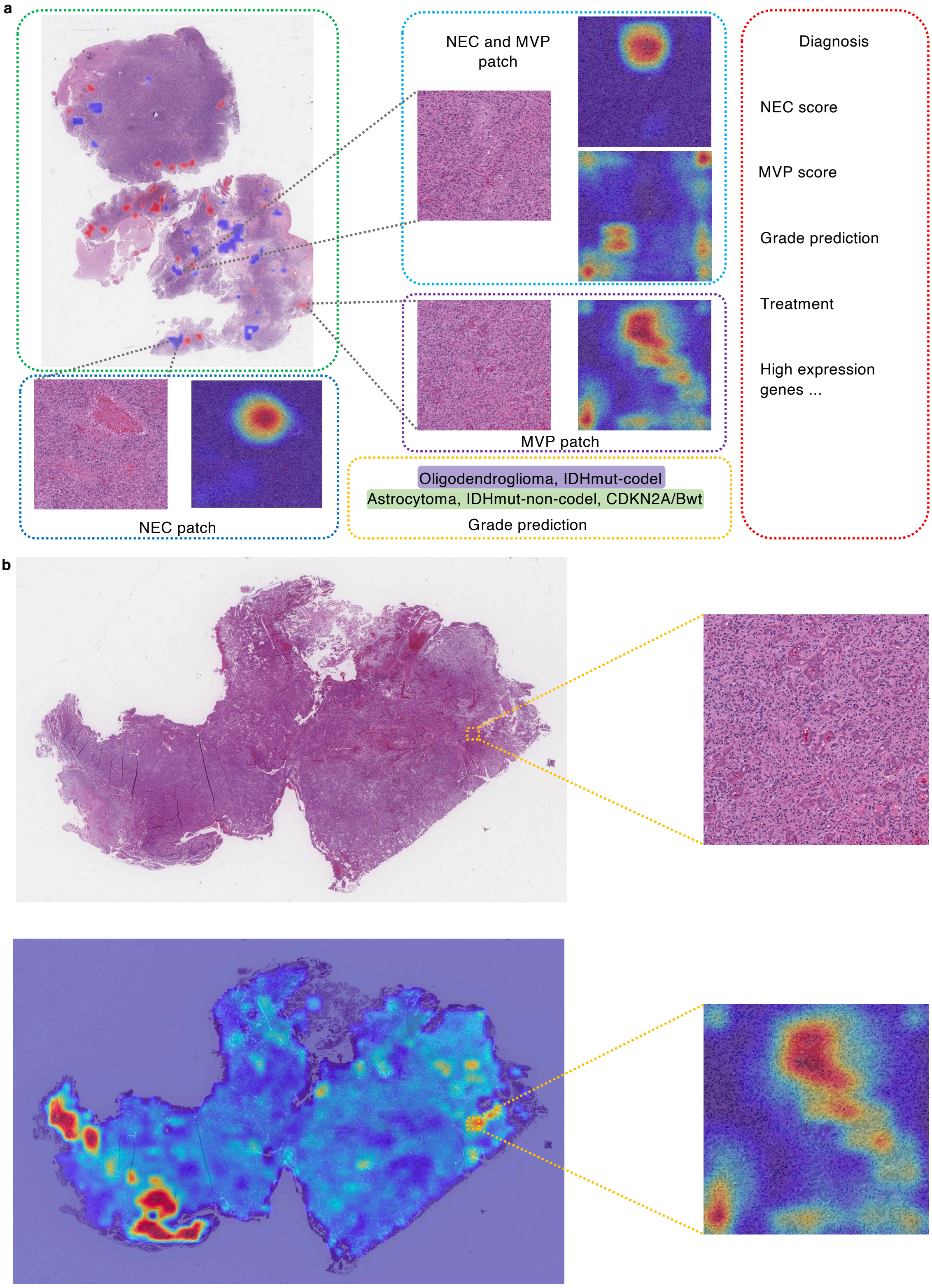
