## Supplemental Mauscript, Figures and Table for "Deep learning-based identification of necrosis and microvascular proliferation in adult diffuse gliomas from whole-slide images": Supplementary.docx

**List of Supplementary Documents:**

**SMa. 1 refers to**

**Supplementary Materials 1: Molecular features and subgroups**

The molecular features of 1,122 TCGA patients included IDH mutation, 1p/19q co-deletion, TERT promoter mutation, EGFR gene amplification, chromosome 7 gain combined with chromosome 10 loss and CKDN2A/CDKN2B homozygous deletion. The patients could be separated into 4 groups, which were Oligodendroglioma (IDH-mutant and 1p/19q-codeleted, WHO Grade 2/3), Astrocytoma (IDH-mutant, 1p/19q-non-codeleted and CDKN2A/B wildtype, WHO Grade 2/3), Astrocytoma (IDH-mutant, 1p/19q-non-codeleted and CDKN2A/B HD, WHO Grade 4), Glioblastoma (IDH wild type, WHO Grade 4).

**SMe. 1 refers to**

### **Supplementary Methods 1: Model selection for patch level**

DenseNet is a kind of network architecture that connects each layer to every other layer in a feed-forward fashion that differs from traditional convolutional networks which only connect layers sequentially. DenseNet's key design is its dense connectivity pattern, where each layer is connected to every other layer, improving the network's efficiency in feature reuse and reducing the vanishing gradient problem. EfficientNet is a kind of network architecture that uses a compound coefficient to uniformly scale all dimensions. Unlike conventional practice that arbitrary scales these factors, the EfficientNet scaling method uniformly scales network width, depth, and resolution with a set of fixed scaling coefficients. The base EfficientNet network is based on the inverted bottleneck residual blocks of MobileNetV2, in addition to squeeze-and-excitation blocks. EfficientNet is suitable for a wide range of applications, including those with limited computational resources. It enhances the representational capability of the network through the incorporation of Squeeze-and-Excitation blocks, which employ an attention mechanism. ResNet is a kind of network architecture that employs a technique called "residual mapping". Instead of hoping that every few stacked layers directly fit a desired underlying mapping, the residual network explicitly lets these layers fit a residual mapping. The skip connections in ResNet can effectively prevent gradient vanishing, thereby enhancing feature representation.

**SMe. 2 refers to**

### **Supplementary Methods 2: Transfer learning on patch-level models**

The last layer of the networks was reinitialized to suit our binary classification tasks. Both the NEC and MVP patch-level models were trained for 100 epochs with a batch size 8, employing the Adam optimizer with an initial learning rate of 1E-04, weight decay, cosine annealing learning rate scheduling, and a cross-entropy loss function. Prior to training, patches were resized to 512 $\times$ 512 pixels before being fed into the network for processing, and data augmentation techniques such as random rotation, random shift, shearing, zooming, and flipping were applied.

**SMe. 3 refers to**

**Supplementary Methods 3: Patient-level models training and validation**

We employed 5-fold cross validation on the datasets labeled by TCGA pathology reports. The NEC patient-level model was trained for 200 epochs with a batch size 4, utilizing the Adam optimizer with an initial learning rate of 2E-04, weight decay, a 50 steps learning rate schedule, and a cross-entropy loss function. The MVP patient-level model was trained for 200 epochs with a batch size 16, utilizing the Adam optimizer with an initial learning rate of 1E-04, weight decay, a 50 steps learning rate schedule, and a cross-entropy loss function. The patches were sized at 224 $\times$ 224 pixels and served as inputs to the network during training, and no data augmentation was implemented.

**SMe. 4 refers to**

**Supplementary Methods 4: Patients categorization**

Within each cluster, we randomly selected 50 patches from NEC and MVP for manual inspection, respectively. Notably, none of the 50 patches in cluster 2 were identified as PAN, while 38 out of 50 patches in cluster 1 were classified as PAN patches. A similar phenomenon was observed where 5 out of the 50 patches in cluster 2 were identified as GMP, while 42 out of 50 patches in cluster1 were classified as GMP. This included the PAN enriched patient group (defined as having 2 or more PAN patches in one WSI), and other necrosis enriched patient group (defined as having fewer than 2 PAN patches in one WSI). Similarly, patients were classified into the GMP enriched patient group (defined as having 6 or more GMP patches in one WSI), and other MVP enriched patient group (defined as having fewer than 6 GMP patches in one WSI).

**SMe. 5 refers to**

### **Supplementary Methods 5: Differential expression and gene set enrichment analyses**

We conducted differential expression and gene set enrichment analysis (GSEA) utilizing the following R packages: TCGAbiolinks, DESeq2, fgsea, EnhancedVolcano, and the Python package: GSEApy, utilizing the normalized RNA-seq expression data acquired for the TCGA glioma dataset. The Hallmark gene sets were downloaded from the Molecular Signatures Database (MsigDB; version 7.1). Firstly, we employed the TCGAbiolinks to download the RNA-seq expression data from the National Cancer Institute (NCI) Genomic Data Commons (GDC) thorough its GDC Application Programming Interface (API), filtered out the samples without WSI, and obtained 455 samples. Secondly, we selected samples from two groups based on the PAN clustering result: PAN enriched patients (n=42) and other necrosis enriched patients (n=70), and GMP clustering result: GMP enriched patients (n=56) and other MVP enriched patients (n=71). Finally, we ran the DESeq2 and EnhancedVolcano for the differential expression analysis with volcano plot and used fgsea and GSEApy for gene set enrichment analysis (GSEA) with the permutation type set to gene set and the number of permutations set to 1000 for GSEA. Only Hallmark gene sets with a p-value < 0.05 were selected as the functionally enriched biological states or processes.

**Table S1:** Re-grading of the TCGA Low Grade Glioma cohort based on our model and the 2016 and 2021 WHO classification of Tumors of the Central Nervous System.

**Figure S1 to S7 were extended data for main figures**

**Figure S1 refers to**

**Supplementary figure 1 | Data preprocess.** (**a**) Discarding background and generating patches. (**b**) Staining normalization. A standard patch was shown on the left and 3 example patches extracted from 3 various WSIs were re-stained based on normalization algorithm.

**Figure S2 refers to**

**Supplementary figure 2 | Performance of the patch-level deep learning models in training, validation, and test datasets.** (**a**) Overall performance comparison between model and pathologists (reference) in classifying NEC and MVP patches. (**b**) Histogram plots with kernel density estimation (KDE) showing the prediction probability distribution of NEC (upper panel) and MVP (lower panel) patches by deep learning models. (**c**) Performance comparison with pathologists in predicting NEC (MVP) patches in one WSI.

**Figure S3 refers to**

**Supplementary figure 3 | Performance of models in the prediction of training, validation, and test datasets.** (**a-c**) Performance comparison with pathologists in predicting NEC and MVP patches for DenseNet201, EfficientNetB2, and ResNet50 models.

**Figure S4 refers to**

**Supplementary figure 4 | Pipeline for processing pathology reports of TCGA WSIs.** (**a**) Assemble and match patch-level probability predictions and coordinates to form the probability localization maps (size: 224x224) for WSIs. (**b**) Employing optical character recognition (OCR) to extract searchable text documents from pathology reports in PDF format and utilizing fuzzy keyword search for NEC and MVP to acquire patient-level labels for the WSIs. (**c**) Caption content of all the pathology reports. (**d**) Detailed architecture of Probability Localization Network (PLNet). Convolutional layer (Conv), Batch normalization layer (BN), and Rectified linear unit (ReLU).

**Figure S5 refers to**

**Supplementary figure 5 | Survival analysis using the patient-level model.** (**a**) Survival curves based on the most significant p-value for O, IDHmut-codel from **Fig. 5(e)** and (**g**). (**b**) Based on 186 records with CDKN2A/B wildtype from all *A, IDHmut-non-codel* samples in TCGA, and considering the results in **Fig. 5**(**f**) and (**h**), we further subtyped the samples with more significant NEC/MVP. These records have the potential to be of a higher grade. (**c**) Based on 160 records with *O, IDHmut-codel* in TCGA, and considering the results in (**a**), we further subtyped the samples with more significant NEC/MVP. These records have the potential to be of a higher grade.

**Figure S6 refers to**

**Supplementary figure 6 | Hallmark gene set enrichment analysis.** (**a-b**) Gene set enrichment analysis (GSEA) showing the significantly (*P* < 0.05) altered pathways in PAN enriched patient group compared with Other NEC enriched patient group, and GMP enriched patient group compared with Other MVP enriched patient group. (**a**) A positive normalized enrichment score (NES) value indicates enrichment in the PAN, a negative NES indicates enrichment in Other NEC. (**b**) A positive NES value indicates enrichment in the GMP, a negative NES indicates enrichment in Other MVP. (**a-b**) Previous studies investigated that PAN in glioblastoma was associated with hypoxia and GMP in glioblastoma was associated with angiogenesis, which were consistent with our findings. (**c**) GSEA results indicating the highest normalized enrichment scores were observed in the E2F targets pathway for both PAN and GMP enriched patients.

**Figure S7 refers to**

**Supplementary figure 7 | Results combination for generating diagnosis reports and online demonstration.** (**a**) Combining all the NEC and MVP prediction results to generate the diagnosis reports. (**b**) Combining the patch-level predictions to generate the patient-level heatmap for online demonstration.
